# Improving geographical accessibility modeling for operational use by local health actors

**DOI:** 10.1101/2020.03.09.20033100

**Authors:** Felana Angella Ihantamalala, Vincent Herbreteau, Christophe Révillion, Mauricianot Randriamihaja, Jérémy Commins, Tanjona Andréambeloson, Feno H Rafenoarivamalala, Andriamihaja Randrianambinina, Laura F Cordier, Matthew H Bonds, Andres Garchitorena

## Abstract

**Background:** Geographical accessibility to health facilities remains one of the main barriers to access care in rural areas of the developing world. Although methods and tools exist to model geographic accessibility, the lack of basic geographic information prevents their widespread use at the local level for targeted program implementation. The aim of this study was to develop very precise, context-specific estimates of geographic accessibility to care in a rural district of Madagascar to help with the design and implementation of interventions that improve access for remote populations.

**Methods:** We used a participatory approach to map all the paths, residential areas, buildings and rice fields on OpenStreetMap (OSM). We estimated shortest route from every household in the District to the nearest primary health care center (PHC) and community health site (CHS) with the Open Source Routing Machine (OSMR) tool. Then, we used remote sensing methods to obtain a high resolution land cover map, a digital elevation model and rainfall data to model travel speed. Travel speed models were calibrated with field data obtained by GPS tracking in a sample of 168 walking routes. Model results were used to predict travel time to seek care at PHCs and CHSs for all the shortest route estimated earlier. Finally, we integrated geographical accessibility results into an e-health platform developed with R Shiny.

**Results:** We mapped over 100,000 buildings, 23,000 km of footpaths, and 4,925 residential areas throughout Ifanadiana district; this data is freely available on OSM. We found that over three quarters of the population lived more than one hour away from a PHC, and 10-15% lived more than one hour away from a CHS. Moreover, we identified areas in the North and East of the district where the nearest PHC was further than 5 hours away, and vulnerable populations across the district with poor geographical access (>1 hour) to both PHCs and CHSs.

**Conclusion:** Our study demonstrates how to improve geographical accessibility modeling so that results can be context-specific and operationally actionable by local health actors. The importance of such approaches is paramount for achieving universal health coverage in rural areas throughout world.

## Background

In 2018, world leaders celebrated the 40 years since the adoption of the Alma Ata Declaration, which recognized the need to invest in primary care as the key to attaining the goal of “Health for All”. While we have witnessed since then an unprecedented global improvement in health indicators, half of the world’s population continues to lack access to essential health services [1]. Low health care access in rural settings of the developing world is due to a combination of financial, geographical and health system barriers [2–6]. To reduce the impact that weak health systems and user-fees have on health care access, there is a growing consensus around the central importance of sector-wide approaches such as health system strengthening (HSS) and universal health coverage (UHC) [7–9]. An increasingly recognized pillar of many HSS and UHC efforts is the role of community health workers (CHWs) to reduce geographical inequities in access and ensure the delivery of primary care at the community level [10].

Community health strategies are underway in most rural areas of Sub-Saharan Africa, where health infrastructure is sparse and the majority of travel is on foot [10]. Distance and travel time to primary health centers (PHC) in these areas are known drivers of care utilization, showing consistent negative impacts on the use of prenatal, perinatal and obstetric care for women [4, 11–15]; child vaccination coverage and pediatric health utilization [16–19]; voluntary enrolment in health insurances [2]; and rates of diagnosis or treatment for tuberculosis, malaria, and HIV [20–24]. In fact, the use of primary care tends to fall exponentially as distance from facility rises, a phenomenon known as “distance decay” [16, 23, 25–27]. Geographical barriers to care can persist even when facility-based HSS activities are in place, making these approaches insufficient to reach full population coverage of primary care services [28–33]. Therefore, to optimize both facility-based and community-based strategies towards the realization of UHC, a much deeper understanding of geographical accessibility to PHC and CHS is necessary in contexts undergoing HSS efforts.

Geographical access to health care has been previously characterized in other contexts using a variety of methods [13, 34–36], aimed at estimating distance or travel time to reach health facilities for populations [37, 38]. A common approach consists of estimating travel time through population surveys, but this method is resource-intensive and prone to biases related to subjective measures of time [23, 26, 34]. An alternative approach is the use of geomatics with available geographic information [39–41], either estimating Euclidean distance (“as the crow flies”) or using more precise algorithms that account for terrain characteristics and road networks through friction surfaces [35, 39, 42, 43]. To increase the adoption of geographic accessibility analyses into health planning, the WHO integrated those algorithms into an easy to use, freely available tool (“AccessMod”) that includes multiple functionalities [44, 45]. However, methods that rely on friction surfaces represent only a “best guess” of the routes people use in areas with poor road infrastructure and of the speed at which people travel. For geographical accessibility analyses to be precise enough for local use by program managers and health workers, reliable information on footpath networks needs to be combined with context-specific estimates of travel speed, and integrated via e-health tools. This can inform the design and implementation of geographically targeted interventions that balance facility-based, community health, and outreach strategies in order to maximize population access to primary care.

Such approaches are particularly needed in Madagascar, a country with one of the least funded health systems in the world [46]. In 2014, Madagascar had less than 3 clinicians (doctors, nurses and midwives) per 10,000 people [47], with a lower concentration in rural areas, where over three quarters of the population live [48]. Access to health care is particularly low for populations living more than 5 kilometers away from a PHC, putting them at higher risk for early childhood mortality [49, 50]. In 2014, the Madagascar’s Ministry of Health (MoH) and the nongovernmental organization PIVOT partnered to strengthen the public health system in the rural district of Ifanadiana, with the aim of attaining universal health coverage and set a model for the country. Despite rapid improvements observed in accessibility and health conditions [51, 52], initial analyses suggested that health gains were concentrated in close proximity to health centers [52]. Here, we aimed to develop very precise, context-specific estimates of geographic accessibility to care in Ifanadiana district to help with the design and implementation of interventions that improve access for remote populations. We mapped all buildings and footpaths in the district to accurately estimate the shortest route to health facilities, and we parametrized travel time estimations with hundreds of hours of fieldwork and remote sensing analyses. We integrated all this information into accessible e-health tools for use by PIVOT, MoH and local partners.

## Materials and Methods

### Study area

The study area is Ifanadiana, a rural health district located 444 km southeast of Antananarivo. The district has an area of 3,975 sq. km and is characterized by a mountainous landscape. The district’s health system is comprised of one hospital (CHRD II), 21 PHC facilities and 195 CHS where CHWs provide consultations for children under 5 years and reproductive women; these may be their homes or a designated structure. There is only one paved road crossing the district from West to East (national road RN25) and through Ranomafana National Park. Additionally, there are two non-paved axes connecting the main towns in the North and South of the District, which are partly accessible by 4WD vehicles or all-terrain motorcycles. Most villages in the District are connected to each other by small paths only accessible by foot. High rates of extreme poverty, geographical barriers, and unreliable health services were associated with very limited access to health care in the district in 2014, which was substantially lower than average estimates for Madagascar [50, 53]. Since then, the NGO PIVOT has worked in Ifanadiana in partnership with the Ministry of Health to create a “model district”, so that the experience in this district can help improve national strategies and health policies throughout the country. The intervention included the removal of most point-of-service payments as well as improved facility readiness and clinical programs at all levels of care (*i*.*e*. hospital, health centers, and community health). In particular, Ifanadiana is one of the first districts to officially pilot the national policy on Universal Health Coverage, which aims to ensure access to quality healthcare for all through strengthened health systems and a reduction of point-of-care fees. Moreover, PIVOT is piloting alternative, professionalized, models of community health through enhanced supervision by certified nurses, building infrastructure for CHSs in partnership with local communities, and implementing proactive community case management. The work described in this study was in support of these two major initiatives.

### Data collection

### Participatory mapping with OpenStreetMap

Detailed, freely available data on footpath networks and villages in rural areas of the developing world are necessary to obtain precise routes for accessing care, but this information is largely absent. To fill this gap, we carried out photo-interpretation using very high spatial resolution satellite images in OpenStreetMap (OSM), a collaborative mapping project with tools for drawing roads, houses, and land use contours among others [54]. For this, we collaborated with the Humanitarian OpenStreetMap Team (HOTOSM) [55], an organization that promotes collaborative mapping projects on the OpenStreetMap platform for humanitarian purposes through a dedicated interface and network to a large online community. The district was divided into 3,508 tasks of 1 sq. km each. Mapping of each task was done in a two-stage process. First, one or several individuals mapped all paths, roads, buildings, and residential areas (defined as groups of 4 or more buildings) within a particular task. This was done on OSM using Digital Globe Standard imagery for background (30 – 60 cm spatial resolution), and Bing maps imagery (up to 30 cm spatial resolution) as backup when cloud cover in Digital Globe images prevented their use for mapping. After the task was marked as mapped, it was available for validation by a separate person. The validation stage, which uses the same tools as the mapping stage, allowed making all necessary corrections of each task in order to ensure the consistency and quality of the mapping. After the completion of this mapping in HOTOSM, we carried out an additional mapping of the hydrographic network (streams and rivers) and rice fields, following the same protocol. OSM mapping of Ifanadiana district was achieved in 8 months with the collaboration of 103 participants. To increase participation in the mapping project, we organized 5 “mapping parties” with local universities and OSM groups in both Madagascar and La Reunion. Despite it being a collaborative project and published in the HOTOSM Task Manager, we had few spontaneous contributions and 5 people from our research team mapped 73.6% of the overall project. The geographic data mapped in OSM is now freely accessible to any user and can be queried on QGIS [56] via the QuickOSM plugin, which we used here for retrieving the data for our analyses.

### Recording travel time on the field

Most travel in Ifanadiana district is done by foot due the minimal transportation infrastructure and steep terrain. To obtain context-specific estimates of travel speed by foot according to terrain characteristics, we recorded GPS data from 168 walking routes across 10 out of the 15 communes of Ifanadiana district between September 2018 and April 2019 (Additional file 1). We collected two types of routes: 1) routes from field expeditions of PIVOT’s community team staff during CHW supervisions, and 2) routes specifically recorded for this project to obtain a larger sample size and wider representation of terrain characteristics, collected by representatives of the PIVOT research team and by the local population (Additional file 1). We recorded these tracks using Samsung Tab A10 tablets and the Android app “OsmAnd” version 3.0.2 [57]. OsmAnd is a free map and navigation app based on the OSM database. For each trip, we recorded via OsmAnd the GPS location, time and altitude every 10 seconds.

#### Satellite imagery and remote sensing

We complemented the mapping work in OSM, which provides some elements of land use, with remote sensing analyses of satellite images to identify forests, water bodies and savanna land uses. For this, we used free Sentinel-2 images (level-2A) from August 18, 2018, which were orthorectified, provided Top Of Canopy (TOC) reflectance, and had a 10m spatial resolution. We used the Dzetsaka plugin for semi-automatic classification in QGIS [58]. First, we manually outlined over fifty polygons representing regions of interest (ROI) for each of the three classes (forests, water bodies and savanna). We then ran random forest algorithms, which have good performance in the classification of remotely sensed data with good accuracy [59]. The random forest model calculates a response variable by creating many different decision trees and then allocating each multi-layered pixel down each decision tree. The response is then determined by evaluating the responses from all trees. The class that is predicted the most is the class that is assigned to the object. Forty percent of the image’s pixels were used for validation. Second, we used the model to predict values for the whole satellite image in order to obtain a classified image. The last step of the process consisted of a post-classification, where smaller clusters of areas under 10,000 sq. m were removed and replaced by the pixel value of the largest neighbor polygon. This process was done to improve the quality of the classification product. Finally, we merged these supervised classifications with the two thematic classes obtained earlier through OSM (residential areas and rice fields). We validated the land use map by recording 62 control points on the field during four expeditions across the district and we completed these observations by identifying 254 points through Google Earth. We finally computed a confusion matrix to compare observed and classified values by class.

In addition to land cover, we obtained elevation and precipitation data from remotely sensed data. We downloaded the Shuttle Radar Topography Mission (SRTM) Digital Elevation Model (DEM) from the United States Geological Survey (USGS, [60]), which gives elevation with a 30 m ground resolution. We also acquired precipitation estimates from NASA Prediction of Worldwide Energy Resources (POWER) Project [61], with a spatial resolution of 0.5 * 0.5 degree.

#### Estimation of shortest path distance

Once the mapping of all buildings and footpaths was finalized, we used the OSRM software [62] to calculate the distance of the shortest path between each building and the closest health facility, via the R software package “osrm” (Figure 1). OSRM uses the Dijkstra’s routing Algorithm, which searches iteratively the shortest path from a single node to the destination node in a network. For each building, the shortest path distance to two health facilities were calculated: to the closest PHC and to the closest CHS. When the precise location of a CHS was unknown, we assumed that it was located in the main village of the Fokontany (“*chef lieu*”), as indicated in national policies for community health. In addition to the distance values, the actual shortest path was saved as a vector file (shapefile format) for use in travel time estimations (next section). Finally, we interpolated the distance values in the whole district using kriging methods available in ArcGIS to improve visualization of results.

**Figure 1:**
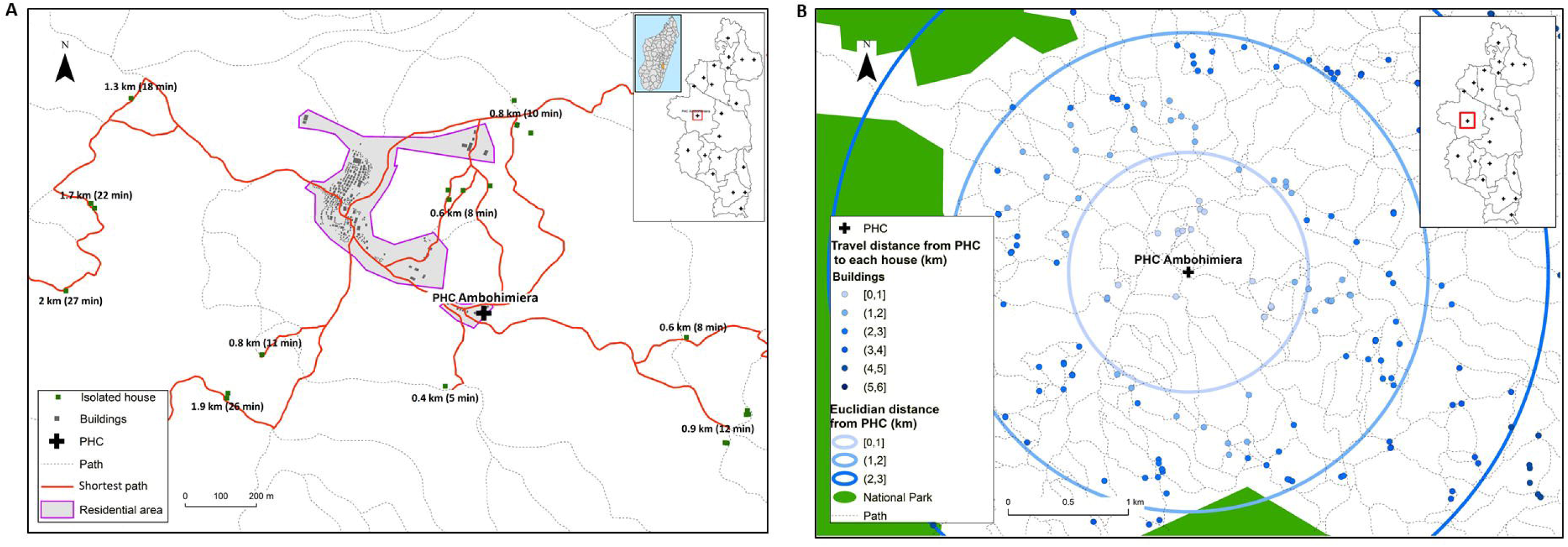
Estimation of the shortest paths from a building to join the PHC. A) Shows an illustrative example of shortest paths obtained thought OSRM, with building values for travel distance and time to reach one of the district’s PHC B) Shows how the travel distance calculated by OSRM improves on typical Euclidian distance estimations, providing more realistic and accurate values by using the footpath network.

#### Estimation of travel speed and time to seek treatment

To obtain precise and context-specific estimates of time to seek treatment, we studied the geographic and climatic factors associated with travel speed in the sample of 168 walking routes collected on the field. For this, travel speed between each pair of points within a GPS track was estimated for each track using the time and GPS location at which each point was taken. Then, using the raster data from the land use classification, DEM and rainfall, we intersected these data with the 168 walking routes. As a result, between each pair of points (N= 57,719) we obtained values for the following explanatory variables: degree of slope, cumulative distance since the beginning of the track, precipitation and land use.

We modelled the impact of each geographic and climatic factor on travel speed using additive models that included a random intercept for each individual track. First, exploratory and univariate analyses were carried out to understand the relationship between each variable and travel speed, including linear and non-linear relationships for slope as well as categorical and numerical variables for land use and cumulative distance (Additional file 2). Cumulative distance since the beginning of the track was categorized following exploratory analysis into 2 groups (0 to 13 km and 13 to 30 km) to reflect the reduction in speed after substantial walking. The land use was converted into a categorical variable that represented the predominant thematic class between each pair points, and a category " mixed " was added when the predominant class represented less than 50%. Slope was included as a non-linear smooth in the additive model. Each explanatory variable with a p-value under 0.1 was included in the multivariate analysis, and model fit was estimated via AIC (Akaike information criterion). The model with the lowest AIC was selected. Model validation was carried out to check for normality, homogeneity and independence of residuals.

Using travel speed estimates from the fixed effects of the final multivariate model, travel time was predicted for each of the 41,426 routes obtained through OSRM (two per isolated building or residential area, one to the closest PHC and one to the closest CHS). For this, we divided these routes into 100 m segments and intersected each segment with DEM and land use to obtain the same set of explanatory variables. Since rainfall affects travel speed and varies from day to day, for each route we provided a prediction for a scenario without rainfall (minimum time) and with the maximum amount of rainfall recorded during fieldwork (maximum time). As with travel distance, we used kriging methods to interpolate the values of travel for the entire district to improve visualization.

#### Development of an e-health tool with R Shiny

We developed an online app to facilitate use of the data and results from the study by local health staff. It consists of a website interface that builds on the estimation methods for distance and travel time in Ifanadiana district presented here, to make the results flexible and easily accessible by program managers and health workers (in French). We used the package Shiny [63] for R statistical software. This app is hosted at the PIVOT dashboard website (http://research.pivot-dashboard.org:3838/) for both private and public use.

## Results

### Mapping

A total of 106,171 buildings were mapped. Of these, 65.6% were located in one of the 4,925 residential areas whereas 34.4% were isolated houses. The size of residential areas ranged from 4 to 870 buildings, with an average size of 14. Moreover, 23,726 km of footpaths were mapped, which represent 99.1% of the road network in Ifanadiana district. Only 0.3% of the road network (62 km) are paved secondary roads. To expand available data, we also mapped on OSM the name and location of 707 villages, 21 PHCs (2 of which were recently built), 37 CHSs built by PIVOT. This district GIS dataset was substantially more exhaustive than what is available through OSM for the rest of districts in the Vatovavy Fitovinany region, with an area above 16,788 sq km (Table 1).

**Table 1:** Comparison of geographic information available on OSM in Ifanadiana district following mapping and the average geographic information available in the other districts of Vatovavy-Fitovinany Region.

|  | Ifanadiana |  |  |  | Distric average<sup>1</sup> for the rest of Vatovavy- Fitovinany Region |  |  |  |
| --- | --- | --- | --- | --- | --- | --- | --- | --- |
|  | n | (%) | Length (km) | Area (ha) | n | (%) | Length (km) | Area (ha) |
| **Residential area** | 4,925 |  |  |  | 223 |  |  |  |
| [0,20] | 4241 | 86.11% |  |  | 206 | 92.38% |  |  |
| (20,40] | 371 | 7.53% |  |  | 9 | 4.03% |  |  |
| (40,50] | 58 | 1.18% |  |  | 1 | 0.45% |  |  |
| (50,100] | 177 | 3.59% |  |  | 4 | 1.19% |  |  |
| >100 | 78 | 1.58% |  |  | 3 | 1.35% |  |  |
| **Buildings** | 106,171 |  |  |  | 6,723 |  |  |  |
| Isolated house | 36,539 | 34.42% |  |  | 5,086 | 75.65% |  |  |
| On residential area | 69,632 | 65.58% |  |  | 1,637 | 24.35% |  |  |
| **Rice field** | 17,446 |  |  | 13,436 | 201 |  |  | 297 |
| **Road networks** |  |  |  |  |  |  |  |  |
| Path |  |  | 23,726 |  |  |  | 400 |  |
| Secondary road |  |  | 62 |  |  |  | 68 |  |
| Tertiary road |  |  | 130 |  |  |  | 45 |  |

<sup>1</sup>The average here represents the sum of the number of residential areas, buildings, rice fields and the length of road networks in the other 5 districts of the region (Manakara, Mananjary, Ikongo, Vohipeno, Nosy-Varika) divided by 5.

The remote sensing analysis allowed to map 850 sq km of forest, 12 sq km of water bodies and 2,967 sq km of savanna (21.4%, 0.30 and 74.6 % of the district surface, respectively) at a 10m resolution (Additional File 3). The kappa index for the land use classification was 0.95 [64].

### Shortest path distance to health facilities

Travel distance to reach the nearest PHC varies from 0 to 27 km with a district average of 8 km (Additional files 4 and 5). The most remote areas in terms of PHC accessibility are located in the North (Fasintsara), South-West (Ranomafana) and East of the district (Tsaratanana), where populations live further than 20 km from the nearest PHC (Figure 2). More than two thirds of the district population live further than 5 km from the closest PHC, with 44.43% living between 5 and 10 km, and 24.67% living between 10 and 20 km (Table 2). Only 8.54% live under 2 km from a PHC. Travel distance to reach the nearest CHS varies from 0 to 13 km, with a district average of 2.69 km (Additional files 4 and 5). In several areas across the district, populations live between 4 and 6 km to the closest CHS (Figure 3). Overall, less than 5% of the population resides more than 5 km away from a CHS, but half (52.2%) live between 2 and 5 km. Only 43.75% live under 2 km from a CHS (Table 3).

**Table 2:** Distribution of the population in Ifanadiana according to their distance to the nearest health facility

| Health facility type | Travel distance (km) | Population (%) |
| --- | --- | --- |
| PHC | [0,1] | 5.85 |
|  | (1,2] | 2.69 |
|  | (2,5] | 21.02 |
|  | (5,10] | 44.43 |
|  | (10,20] | 24.67 |
|  | (20,30] | 1.34 |
| CHS | [0,1] | 22.07 |
|  | (1,2] | 21.68 |
|  | (2,5] | 52.2 |
|  | (5,10] | 3.98 |
|  | (10,20] | 0.08 |

**Table 3:**
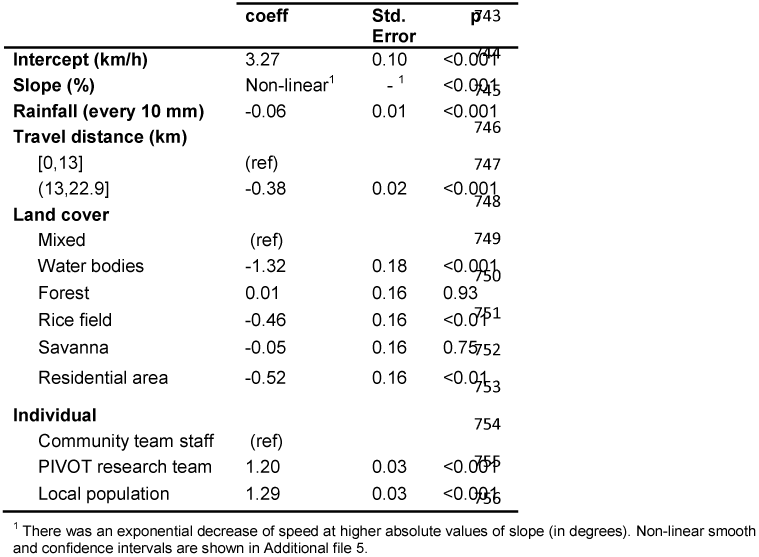
Multivariate analysis of local factors affecting travel speed by foot in Ifanadiana, (linear additive model, with individual track as random intercept)

**Figure 2:**
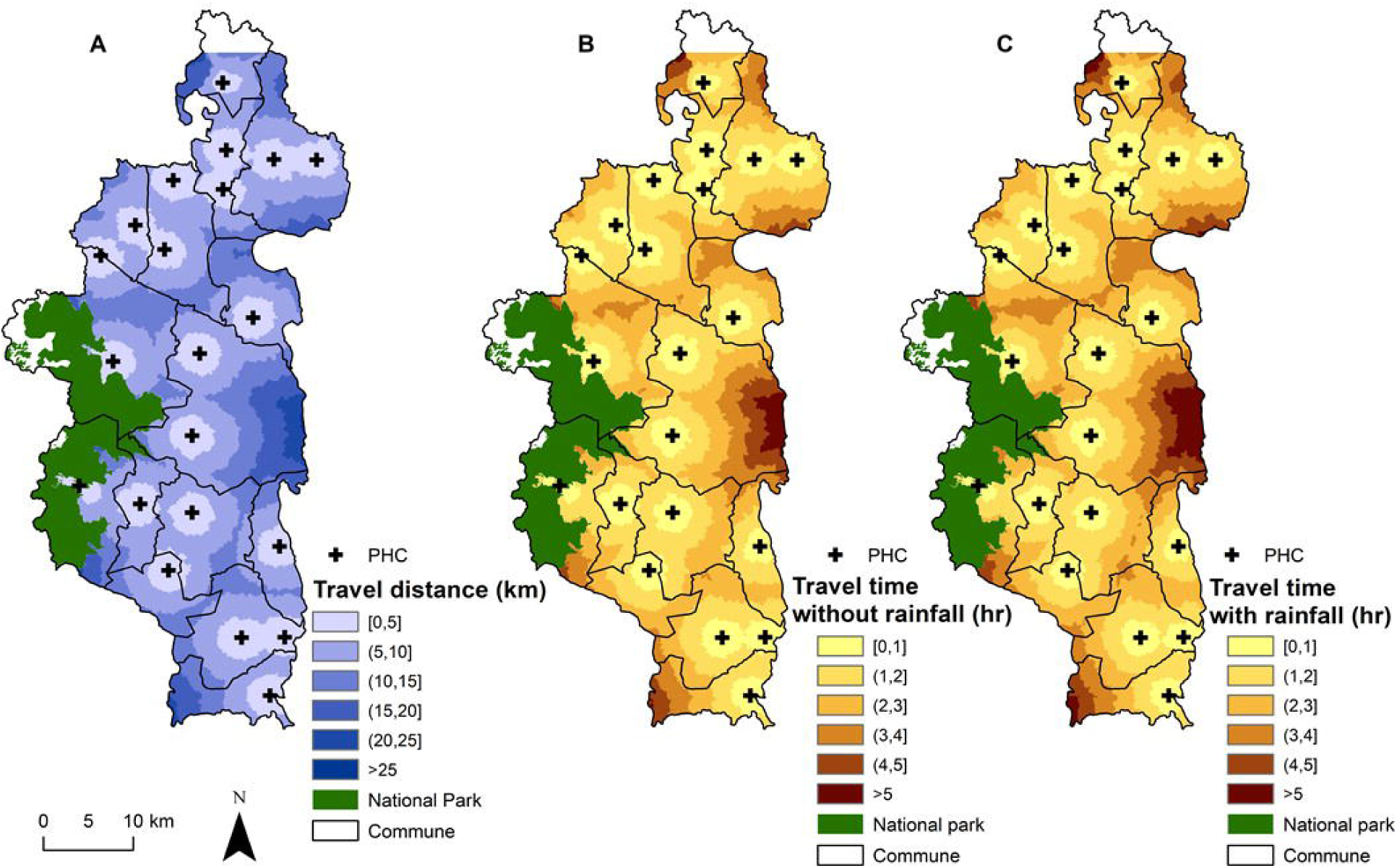
Interpolated distance and travel time between each household and PHCs. A) Spatial variation in the distance to join the nearest PHC, with shades of blue representing 6 distance classes: 0-5, 5-10, 10-15, 15-20, 20-25 and >25 km. B-C) Travel time without and with rainfall to join the nearest PHC, with shades of brown representing 6 time classes: 0-1, 1-2, 2-3, 3-4, 4-5 and >5 hrs.

**Figure 3:**
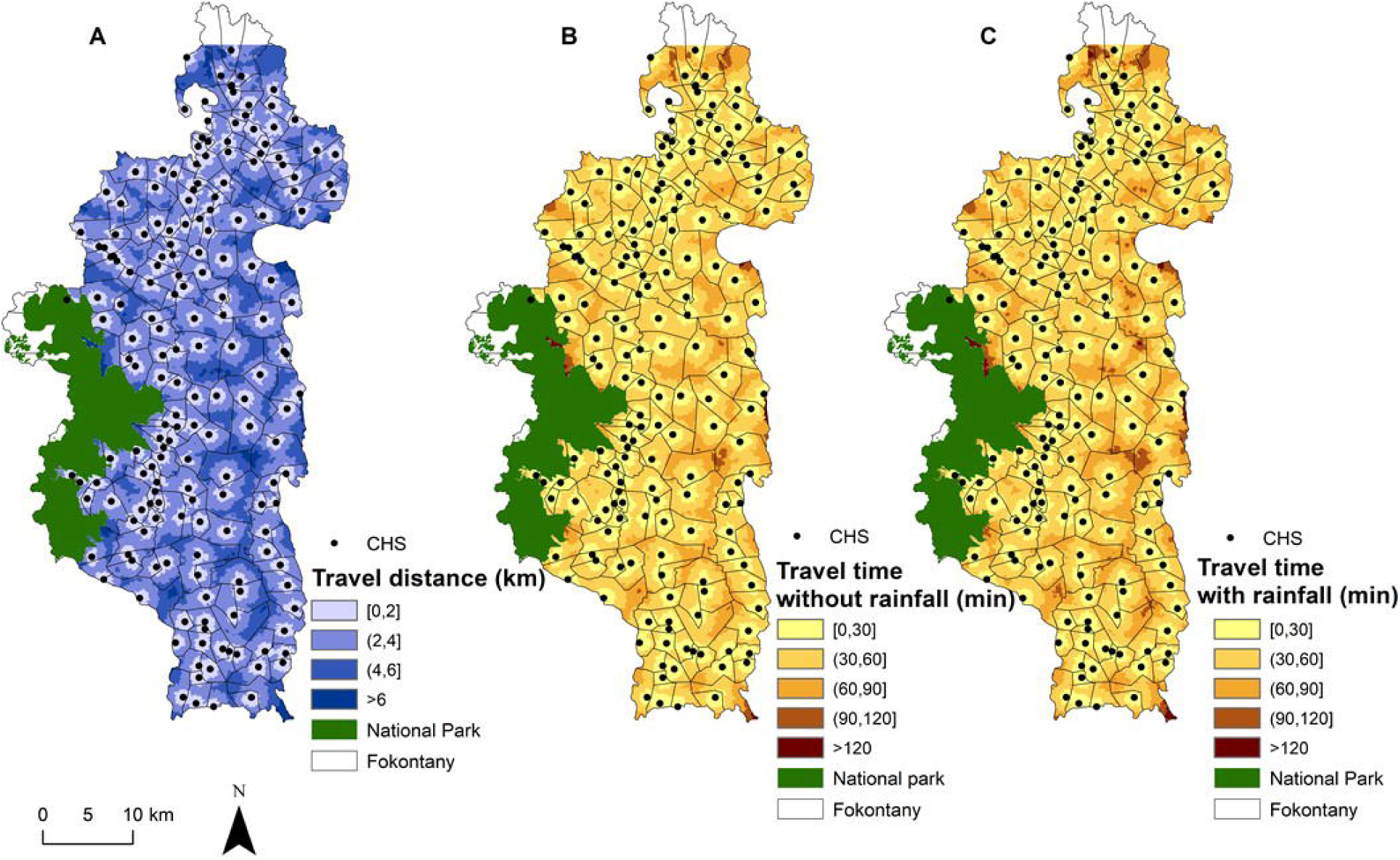
Interpolated distance and travel time between each household and CHSs. A) Spatial variation in the distance to join the nearest CHS, with shades of blue representing 4 distance classes: 0-2, 2-4, 4-6 and >6 km. B-C) Travel time without and with rainfall to join the nearest CHS, with shades of brown representing 5 time classes: 0-30, 30-60, 60-90, 90-120 and >120 mins.

### Time to seek care under different climatic conditions

Statistical analyses from a sample of 168 walking routes in Ifanadiana District (871 km, Additional file 2) allowed us to identify the most relevant determinants of travel speed for local populations and health workers. Overall, there was wide variation in speed between individual tracks, with 39.45% of the variation explained by the random effects. Travel speed was reduced by 0.38 km/h after the first 13 km. In terms of terrain characteristics, slope was the most important determinant of travel speed, which decreased exponentially with absolute values of slope (whether positive or negative, see Additional file 6). Walking through a water body, a rice field, or a residential area were associated with slower travel speeds, whereas walking through savanna, forest or a mixed land use were associated with faster speed. Finally, rainfall had a small negative effect on travel speed, where an increase in 10 mm of rainfall was associated with a reduction of 0.06 km/h in speed.

Using predictions of travel speed for the 41,426 routes obtained through OSRM we found that time to seek care at a PHC varies between 0.49 minutes and 7 hours (one way) under dry conditions (average of 111 minutes) and between 0.51 minutes and 7.5 hours during rainy conditions (average 121 minutes; Additional files 3 and 4). The time difference between rainy and dry conditions was most relevant (over 20 minutes) for areas located further than 15 km to the PHC. Overall, only 21.6% of the population can join a PHC within an hour under rainy conditions and 58.1% within two hours. This percentage increases under dry conditions to 24.3% of population being able to reach the PHC within an hour and 63.2% within two hours.

Travel time to reach a CHS varies between 0 and 150 minutes under dry conditions and up to 165 minutes under rainy conditions (Additional files 3 and 4). For areas located further than 5 km from a CHS, the difference between dry and rainy conditions can be of 15 minutes. Overall, 40% of the population lives within 30 minutes from a CHS, and 85.12% lives within 1 hour under rainy conditions (Table 4). This percentage increases under dry conditions to 44% and 89.83% for populations living within 30 min and 1h to a CHS respectively. Small areas all across Ifanadiana had populations that lived further than 1h from a CHS (Figure 3), and most of these populations lived further than 1h from any type of primary health facility, whether a PHC or a CHS (Figure 4).

**Table 4:** Distribution of the population in Ifanadiana according to their travel time to the nearest health facility

| Health facility type | Travel time (minutes) | Population (%) |  |
| --- | --- | --- | --- |
|  |  | If time estimation without rainfall | If time estimation with rainfall |
| PHC | [0,30] | 9.76 | 8.85 |
|  | (30,60] | 14.49 | 12.77 |
|  | (60,120] | 38.90 | 36.5 |
|  | (120 ,180] | 25.20 | 26.19 |
|  | (180,240] | 7.97 | 10.54 |
|  | >240 | 3.66 | 5.14 |
| CHS | [0,30] | 44.24 | 39.98 |
|  | (30,60] | 45.59 | 45.14 |
|  | (60,120] | 9.95 | 14.59 |
|  | (120 ,180] | 1.21 | 0.27 |
|  | (180,240] |  | 0.03 |

**Figure 4:**
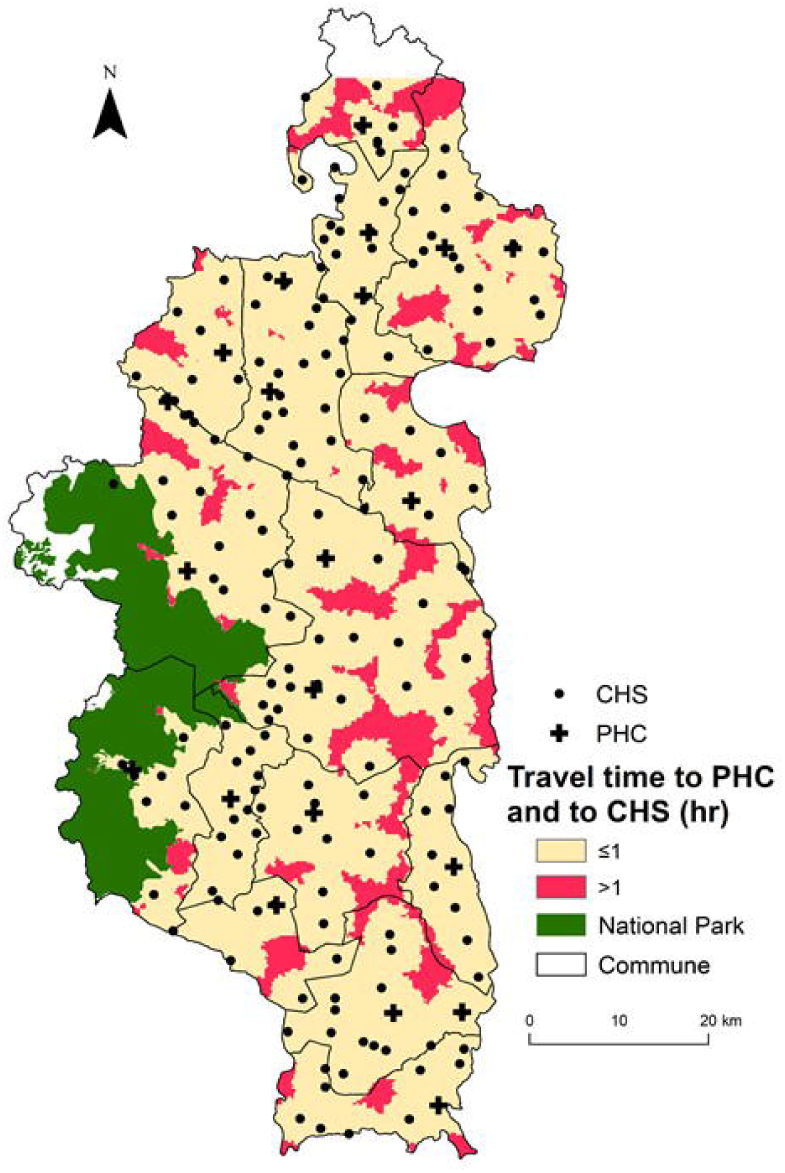
Distribution of vulnerable populations with poor geographic access to both PHC and CHS. It shows the spatial variation in the travel time to reach any type of primary care facility (both PHC and CHS). Areas less than one hour away are shown in light brown and those more than one hour away in red.

### E-health tools for geographical accessibility to care

The web interface created for health workers and program managers (Figure 5) allows visualization of key information for the planning and implementation of health programs such as 1) areas with the lowest accessibility to health care based on shortest path distance and travel time, 2) the percentage of the population in each commune and fokontany that are at a particular distance and travel time to the nearest health facility, 3) a tool to estimate shortest path routes for field expeditions and community health work with either a satellite or OSM background, and 4) the geographic distribution of residential areas and isolated households that are within a certain distance or time from a selected health facility. In addition to the web interface, which can be accessed by phone or computer but requires an internet connection, field workers can get accurate directions without access to the internet through the Android app “OsmAnd”, as the full footpath network and all residential areas have been mapped on OSM.

**Figure 5:**
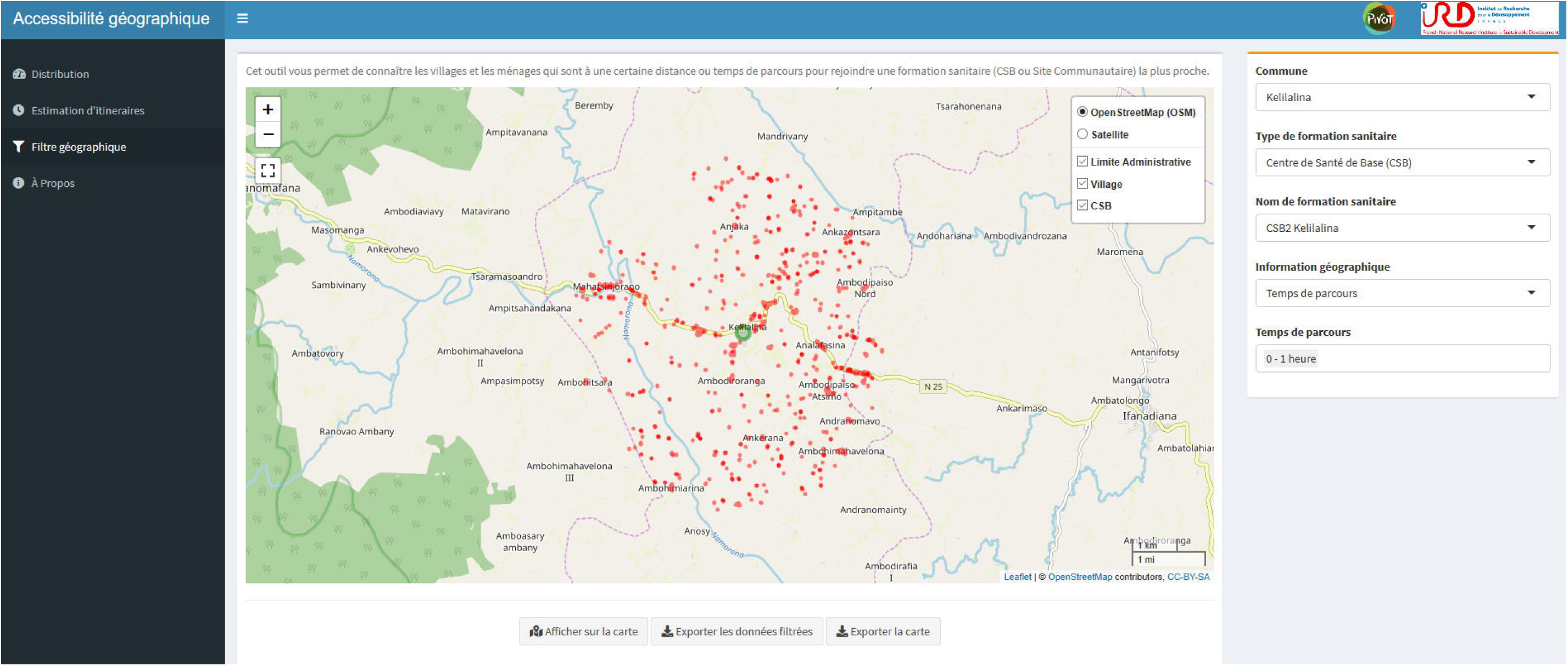
Shiny app for operational use by local health actors. Illustrative example of the interface, showing in a map all the residential areas and isolated houses (red polygons) located between 0 and 1 hour from the PHC (green circle).

## Discussion

Despite a renewed commitment in recent years to UHC and community health as ways to increase financial and geographical access to primary care, half of the world’s population continues to lack access to essential health services [1]. Distance to health facilities remains one of the main barriers to accessing care in rural areas of the developing world, where transport infrastructure is deficient, population density is low and health facilities are sparse. Although methods and tools exist to model geographic accessibility, there is a critical lack of basic geographic information that prevents the widespread use of these tools to optimize the implementation of health programs at the local level. Here, we show how the combination of participatory mapping, fieldwork and remote sensing can provide very precise estimates of distance and time to seek treatment at health facilities that can be used to inform health system design and policy, improve management, and support health workers. We found that over three quarters of the population in Ifanadiana district lived more than one hour from a PHC. Moreover, we identified areas in the North and East of the district where the nearest PHC was further than 5 hours away, and vulnerable communities in most parts of the district with poor geographical access (over 1 hour travel) to both PHCs and CHSs.

A travel time of one or two hours to health services is a generally accepted threshold of poor accessibility to health services, delaying or preventing health seeking behaviors that can have severe consequences [22, 26, 39, 65, 66]. We found that three quarters of the population in Ifanadiana district (76%) lived more than 1h from a PHC and forty percent lived further than 2h, with a larger percentage in the rainy season. This is significantly higher than previous estimates of access to secondary care in sub-Saharan Africa, where only 11-27% were found to live further than 1h from a hospital with surgical capabilities depending on the region [66]. National and sub-national level studies focusing on Emergency Obstetric and Neonatal Care (EmONC) found slightly higher results, with 30% of the women in two regions of Ethiopia [39] and 34% in Ghana living further than 2h from EmONC facilities [26]. Thus, the proportion of the population that potentially lacks access to primary care at PHC in Ifanadiana is comparable to the proportion lacking access to secondary care in other low resource African settings. In addition, because community health is considered a solution for reducing geographical inequities in access to care, geographical accessibility to community health workers is rarely studied. Here we show that over half of the population in Ifanadiana had to walk over 30 minutes to reach a CHS, and 10 to 15% had to walk over 1h depending on season. This suggests that gaps exist even in the geographical coverage of community health programs and opens new questions around how to measure and optimize access to CHSs for rural populations based on a deep understanding of their geographical context.

In this study, we estimated distance to PHC using a mapping of about 23,000 km of footpaths and over 100,000 buildings and parametrized travel time with hundreds of hours of fieldwork, obtaining precise and context-specific accessibility estimates for every community in our district. This approach overcomes many of the challenges that studies of geographical accessibility commonly face. Despite the increasing use of geographical information systems and spatial analyses to better understand health care access in developing countries, there are still important gaps in its application. Studies in high income countries provide accurate estimates of geographical access to services due to the availability of electronic HMIS and GIS data, as well as good transport infrastructure [65, 67–70]. Studies in developing countries typically use either Euclidean distances to obtain a basic measure of distance, or road networks in combination with friction surfaces to obtain estimates of travel time to PHC because the real network of footpaths is rarely available [21, 27, 42]. Given that most travel in rural areas is done by foot, inaccuracies from such models can be quite important [22, 71]. In addition, while travel time can strongly depend on contextual factors, researchers use either constant values for travel speed [20, 26] or different values by geographic feature inputted from other contexts [22]. A 2016 review found that none of the studies on maternal care calibrated spatial accessibility with measured travel time [42]. An alternative to measuring travel distance and time to health facilities is to rely on self-reported answers in surveys, but these are subjective and can be prone to significant bias [23, 26, 34]. Thus, our approach could be replicated in future studies of geographical accessibility to care where very precise estimates of distance and time to seek treatment are required.

This unprecedented level of precision allows for their use locally by decision-makers at the Ministry of Health, program managers and health workers, thanks to the integration with tools that are free, open source, and easily accessible. Mapping on OSM is simple, the information entered is accessible to anyone for download or for use into mobile apps, and the geographic database can be regularly updated or further expanded by the online community. Although in our case most of the work through HOTOSM was done by several mappers involved in the project, this type of participative and collaborative mapping approach can allow for rapid crowdsourced mapping of large geographical areas when the mapping community is heavily mobilized. For instance, following the 2010 earthquake in Haiti, a complete map of Port au Prince was achieved within a month by hundreds of remote mappers through HOTOSM to aid disaster response [72]. Uses of the detailed geographical information available for Ifanadiana (with all the footpaths, hydrographic networks, buildings and rice fields) go beyond its healthcare accessibility applications and could be expanded to other demographic (e.g. population density, migration patterns) or vector-borne disease research projects (e.g. malaria) of public health interest. In addition, Shiny is increasingly popular for the development of interactive web interfaces that take advantage of the statistical power of R software. Applications include among others malaria surveillance [73], health equity [74, 75], precision farming [76], and biological research [77, 78]. In our context, the use of the e-health tools developed here will allow program managers to prioritize and plan community and outreach activities for the most remote populations, while CHWs and field teams will be able to obtain accurate directions anywhere in the district for implementation of such activities.

Our study provides several programmatic insights to improve health care access in Ifanadiana. In the last 3 years, two PHCs have been built in the extreme North and South of Ifanadiana where geographical access was very low, reducing the percentage of population that have to walk 2-5 hours (Additional file 7). Given the current situation, adding 3 new facilities to the 21 existing PHCs in the areas of the district with the lowest geographical access (East, West and Southwest, Figure 2A) would nearly eliminate the need for populations to walk 4 hours or more each way to a PHC. In addition, the national policy for community health establishes that two CHSs should be appointed in every Fokontany, with no regard to the size or spread of the population in the Fokontany. We show that as a result, a non-negligible proportion of the population (∼30,000 people) needs to walk over an hour just to reach a CHS and receive the most basic care. Consequently, PIVOT is piloting the implementation of proactive community care in order to reach everyone despite their distance to a CHS, and is adapting CHW workforce to the demographic and geographic characteristics of each Fokontany. Experiences from other countries suggest that health reforms focused on improving geographical accessibility to primary care can achieve significant results for the most disadvantaged populations [79].

Our study had several limitations. First, dates of the satellite images used for the OSM mapping were not available, which could pose an issue if images did not reflect current conditions, but subsequent fieldwork confirmed that OSM maps (buildings, paths, etc.) were quite accurate. Second, we only considered a model of travel time by foot, which could lead to biases if part of the population travels to PHCs by vehicle. Other studies have indeed estimated travel time by vehicle in settings with good road networks [34, 69], or by both foot and vehicle in low-resource settings [80]. In this context, less than 3% of the population of Ifanadiana has a vehicle, and there is only one paved road (<1% of the road network) where public transportation is available [81]. Third, we infer the proportion of the population living at a particular distance or travel time based on a full mapping of the district’s buildings from high resolution satellite imagery instead of an actual census. Since some of these buildings do not represent inhabited households (e.g. administrative buildings, shops, etc…), this approach could have led to biases if the distribution of non-households was distributed heterogeneously across the district. Fourth, we provide locally parametrized estimations of travel time for Ifanadiana, but our estimates under scenarios with and without rainfall differed only by up to 20 minutes at distances of over 15 km. Although the district has a tropical climate with periods of heavy rains, track GPS recordings of field expeditions were collected during relatively low rainfall periods (up to 47.5 mm per day), which could explain the small impact of rainfall on travel speed. In addition, recordings were done by health workers and community members, so our estimations represent local travel time for healthy individuals. Other groups such as ill individuals, pregnant women or the elderly will likely take longer to reach health facilities, and factors such as break time during a route were not considered, which could be particularly relevant for those who walk for 4 to 8 hours. As a result, values for maximum time to seek treatment at health facilities presented here are probably an underestimation of the real time spent by certain groups or under certain weather conditions.

## Conclusion

In conclusion, this study advances methods to improve geographical accessibility modeling to determine with high precision the distance and travel time between every household and the nearest primary care facilities, so that the results can be context-specific and operationally actionable by local health actors. Integrating the complete mapping of the district with fieldwork and remote sensing analyses into accessible e-health tools may result in better strategic and operational refinement of programs by the MoH and local health partners. The role of such approaches could be transformative to reach the goal of health care access for all in areas where, like in Ifanadiana, the majority of the population face significant challenges to reach primary care facilities and even CHSs.

## Data Availability

Data are available on OpenStreetMap and on the Shinny app described in this study

https://www.openstreetmap.org

http://research.pivot-dashboard.org:3838/

## Abbreviations

CHS: community health site
CHW: community health worker
DEM: digital elevation model
EmONC: emergency obstetric and neonatal care
GPS: global positioning system
HOTOSM: Humanitarian OpenStreetMap Team
HSS: health system strengthening
MoH: Ministry of Health
OSM: OpenStreetMap
OSMR: open source routing machine
PHC: primary health care center
POWER: prediction of worldwide energy resources
SRTM: shuttle radar topography mission
UHC: universal health coverage

## Author’s contributions

FAI, VH, CR, JC, MHB, AG conceived and designed the study. VH, CR, LFC, MHB, AG, advised on the analysis. FAI, MR, TA analyzed the data. FAI, VH, CR, MR, TA, AG contributed to the interpretation of the data. FAI, VH, CR, MR, JC, TA, FHR, AR, LC, MHB, AG drafted the manuscript. All authors read and approved the final manuscript.

## Author’s details

^1^NGO PIVOT, Ranomafana, Madagascar, ^2^Department of Global Health and Social Medicine, Harvard Medical School, Boston, USA, ^3^Institut de Recherche pour le Développement, UMR 228 Espace-Dev (IRD, UA, UG, UM, UR), Phnom Penh, Cambodge, ^4^Université de La Réunion, UMR 228 Espace-Dev (IRD, UA, UG, UM, UR), Saint-Pierre, La Réunion, France, ^5^School of Management and Technological innovation, University of Fianarantsoa, Madagascar, ^6^Ministry of Health in Madagascar, Antananarivo, Madagascar, ^7^ MIVEGEC, Univ. Montpellier, CNRS, IRD, Montpellier, France

### Acknowledgements

We would like to thank all OSM contributors to the cartography of Ifanadiana district, apart from the authors of this article, and in particular the OpenStreetMap Associations of La Réunion and Madagascar for the mapping parties they organized and their individual contributions. We would like to thank also community team staff at PIVOT and community health workers for helping us to collect fieldwork data.

## Availability of data and material

Data are available on OpenStreetMap (https://www.openstreetmap.org) and on the Shinny app described in this study (http://research.pivot-dashboard.org:3838/)

## Consent for publication

All authors approved the manuscript’s submission for publication.

## Ethics approval and consent to participate

Not applicable

